# Estimating number of global importations of COVID-19 from Wuhan, risk of transmission outside mainland China and COVID-19 introduction index between countries outside mainland China

**DOI:** 10.1101/2020.02.17.20024075

**Authors:** Haoyang Sun, Borame L Dickens, Mark I. C. Chen, Alex R Cook, Hannah E Clapham

## Abstract

**Background:** The emergence of a novel coronavirus (SARS-CoV-2) in Wuhan, China in early December 2019 has caused widespread transmission within the country, with over 1,000 deaths reported to date. Other countries have since reported coronavirus disease 2019 (COVID-19) importation from China, with some experiencing local transmission and even case importation from countries outside China. We aim to estimate the number of cases imported from Wuhan to each country or territory outside mainland China, and with these estimates assess the risk of onward local transmission and the relative potential of case importation between countries outside China.

**Methods:** We used the reported number of cases imported from Wuhan and flight data to generate an uncertainty distribution for the estimated number of imported cases from Wuhan to each location outside mainland China. This uncertainty was propagated to quantify the local outbreak risk using a branching process model. A COVID-19 introduction index was derived for each pair of donor and recipient countries, accounting for the local outbreak risk in the donor country and the between-country connectivity.

**Results:** We identified 13 countries or territories outside mainland China that may have under-detected COVID-19 importation from Wuhan, such as Thailand and Indonesia. In addition, 16 countries had a local outbreak risk estimate exceeding 50%, including four outside Asia. The COVID-19 introduction index highlights potential locations outside mainland China from which cases may be imported to each recipient country.

**Conclusions:** As SARS-CoV-2 continues to spread globally, more epicentres may emerge outside China. Hence, it is important for countries to remain alert for the possibilities of viral introduction from other countries outside China, even before local transmission in a source country becomes known.

## Introduction

The emergence of a novel coronavirus (SARS-CoV-2) in Wuhan City, China at the end of 2019, has caused large numbers of cases of coronavirus disease 2019 (COVID-19) and deaths in Wuhan.^1–3^ As of 11^th^ February 2020, there are increasing reports of large scale transmission and numbers of cases in other places in China.^4,5^ In addition, there are now reports of cases in multiple countries outside of China, and limited reports of transmission within countries outside of China.^5^

From January 23^rd^ travel from Wuhan was halted by the Chinese government, and in addition many countries have implemented measures such as airport screening, testing of patients reporting symptoms who have recent travel from China,^5^ quarantining arrivals from Wuhan and/or China or halting travel altogether.

As with many infectious diseases, there is a risk of under reporting of cases, as some people who are infected do not seek care, some who seek care are not diagnosed and, in some settings, those who are diagnosed may not reported. There are also particular issues in an outbreak of a novel pathogen due to difficulties in mobilizing the response and in development of testing capacities, as well as changes over time in the definition of SARS-CoV-2 infection symptoms. There are therefore concerns the reported cases in countries outside of China may be an under-report of what is actually occurring. Previous work has estimated significant under reporting of COVID-19 cases in a number of countries such as Indonesia, Cambodia and Thailand.^6^ A detailed understanding of the geographical distribution of case importation will help to guide resources to places with currently limited capacity to test, and provide support to perform control measures and support for clinical care.

Flight data has been used to determine connectivity between countries and therefore risk of onward transmission from China to other countries. Early in the outbreak, Bogoch et al. listed the countries at most risk of importations given the number of flights from Wuhan. ^7^ Their ranking ultimately followed closely the countries that first reported imported cases, albeit with some countries predicted to report that did not. Since then others have used flight and other data to highlight countries at most risk of importation of SARS-CoV-2 from China. ^8,9^ These papers have then used the infectious disease vulnerability index (IDVI) as an assessment of how at risk a country is to local transmission. ^8,9^

In countries where importations are occurring, it will be important to quantify the risk of onward transmission occurring and the extent of this transmission. Kucharski et al. ^10^ considered this generally given the number of importations using a probabilistic model, and Wu et al estimated a probability of transmission within cities outside China. ^4^ However risk estimates of importations to countries outside China have been focused on the risk of importations from mainland China, with the risk being correlated with the connectivity of places in mainland China to other countries.^7–9^ As transmission increasingly occurs in countries outside mainland China it may also become important to consider the risk of importations to and from other countries. Indeed there have already been cases outside of China, only reporting travel history to countries outside of China, such as from Singapore to the UK,^11^ and from Thailand, Singapore and Japan to South Korea.^12^ In light of this, there is some urgency to assessing the relative risks of onward transmission between countries outside of China.

Therefore in this paper we estimate 1) the number of imported cases globally from Wuhan (using flight data and the currently reported cases), then using our estimates of the number of imported cases and current estimates of R0 for SARS-CoV-2 we estimate, 2) the probability of an outbreak in countries outside mainland China, and finally given this outbreak risk and the flights between countries outside mainland China, we estimate 3) the risk index for importations occurring from and to countries outside mainland China. There is, of course, large uncertainty in these values and we propagate the uncertainty through the estimates.

## Methods

### Data

#### Case data

We used the information on the date, location, and travel history of each reported COVID-19 case, which was synthesized and made publicly available by the nCoV-2019 Data Working Group. ^13^ For each country or territory outside of mainland China, we collated data on the total number of reported COVID-19 cases imported from Wuhan only (not from other places in China), based on the epidemiological data updated as of 7^th^ February—15 days since the Wuhan shutdown (Table S1). For a small number of cases, we were unable to identify the cities from which they were imported. We included these into our data as having originated from Wuhan to avoid producing false positive results when we later on identified countries that may have under-detected SARS-CoV-2 importation from Wuhan.

#### Flight data

We used the monthly number of air ticket bookings during 2017 from the Official Airline Guide ^14^ to approximate the volumes of air passengers for each origin-destination route.

### Statistical analyses

For each country or territory outside of mainland China (denoted by *i*), we assumed that the total number of COVID-19 cases imported from Wuhan (*M*_*i*_) followed a Poisson distribution with rate parameter proportional to the number of air travelers from Wuhan during January 2020 (*υ_WH→і,Jan_*), with an unknown coefficient *λ*_0_ to be estimated from data (more detail in supporting information):

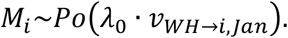

To date, the reported total number of imported cases from Wuhan (*X*_*i*_) differ substantially between countries even after adjusting for the volumes of air passengers arriving from Wuhan. We assumed this was due to inter-country variation in case detection and reporting rates. We ranked countries based on the ratio between the reported case count *X*_*i*_ and passenger volume *υ*_*WH→і,Jan*_, and assumed all the imported COVID-19 cases from Wuhan have been successfully detected and reported by countries having the top 10 rankings, to provide a conservative estimate for *λ*_0_ and hence the number of cases imported from Wuhan to the rest of the countries we consider. In the equation below, *Θ* refers to the set of the aforementioned countries having the highest rankings, and the posterior of *λ*_0_ follows a gamma distribution if we impose a uniform prior:

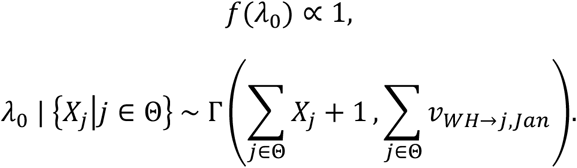

The posterior predictive distribution of the estimated number of COVID-19 cases imported from Wuhan to each country *і ∈ Θ* is thus a Gamma-Poisson mixture. Alternatively, this can be viewed as a negative binomial distribution that models the number of failures before the (∑_*j*∈*Θ*_ *X*_*j*_ + 1)^th^ success in a series of independent and identical Bernoulli trials, each with probability of success ∑_*j*∈*Θ υWH*→*j*,Jan/_(∑_*j*∈*Θ*_ *υ*_*WH→j,Jan*_ + *υ*_*WH→і,Jan*_). Subsequently, we computed the 95% uncertainty interval for the total number of COVID-19 cases imported from Wuhan to each country *і ∈ Θ*. The left tail probability *p*(*M*_*і*_ ≤ *X*_*і*_) can also be used to identify countries that may have under-detected COVID-19 importation from Wuhan.

Next, we propagated the uncertainty in the estimated number of imported COVID-19 cases from Wuhan, and estimated the probability that a local outbreak would occur and sustain for at least three generations (hereinafter referred to as “local outbreak risk”) for each country or territory outside mainland China. We modelled the offspring distribution of each case as a negative binomial distribution, with mean equal to the basic reproduction number estimated by Riou et al. ^15^, and dispersion parameter assumed to be equal to that of SARS-CoV.^16^ Using the first-step analysis, the local outbreak risk can be mathematically derived, where we created two scenarios for each country: (1) only the reported cases imported from Wuhan were immediately isolated, but the rest of the estimated cases were not (main analysis) and (2) immediate isolation of 95% of the estimated imported cases from Wuhan. Here, we assumed that immediately isolated cases were not able to cause any secondary infection throughout their infectious periods, and hence the local outbreak risk estimation was conservative. For the main analysis, we truncated the uncertainty distribution of the imported case count derived earlier using the reported case count, to ensure that the estimated total number of cases imported from Wuhan was always greater than or equal to the reported case count. Countries with a local outbreak risk above 0.5 in our main analysis were named as potential donor countries, and subsequently assessed for their relative potential of exporting SARS-CoV-2 to any recipient country or territory outside mainland China, described as follows.

For each recipient country outside mainland China *r*, we derived a COVID-19 introduction index *γ*_*d*→*r*_ (on a relative scale) that ranks potential donor countries *d* in terms of their viral exportation potential. This was expressed as the product of the probability of travelling from a potential donor country *d* to a recipient country *r* in February, and the local outbreak risk *ω_d_*, where the total population size in each potential donor country (POP_*d*_) was based on the data published by the Socioeconomic Data and Applications Centre ^17^. In addition, we also derived the total COVID-19 introduction index estimate for each recipient country *r* (*γ*_*·→r*_) by summing over all the potential donor countries *d*:

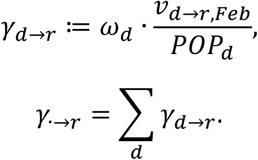

## Results

Outside of mainland China, most countries or territories we estimate as having a large number of COVID-19 cases imported from Wuhan are located in Asia (Figure 1 and Table S1). Outside Asia, we estimate United States, Australia, and United Kingdom as having the highest imported case count estimates (Figure 1 and Table S1). In addition, we also identify countries whose reported number of imported cases from Wuhan is less than the 5^th^ percentile of the posterior predictive distribution of our imported case estimates (Table 1), suggesting under-detected cases imported from Wuhan in these countries or territories. In particular, we estimate that Thailand received 97 (95% CI: 66–136) imported cases from Wuhan—the largest among all the countries analyzed but have only reported ∼20 cases (Table 1). For Indonesia there have not been any reported COVID-19 cases imported from Wuhan, and yet we estimate at least 19 (95% CI: 10–30) imported cases.

**Table 1:** Countries or territories with likely under-detection of COVID-19 cases imported from Wuhan: Reported and estimated case count, and left-tailed P-value.

| Countries or territories | Reported number of COVID-19 cases imported from Wuhan | Estimated number of COVID-19 cases imported from Wuhan | P-value |
| --- | --- | --- | --- |
| Thailand | 20 | 97 (66-136) | < 0.0001 |
| Indonesia | 0 | 19 (10-30) | < 0.0001 |
| Taiwan, China | 10 | 41 (25-61) | < 0.0001 |
| Malaysia | 5 | 27 (15-41) | < 0.0001 |
| Hong Kong, China | 12 | 38 (23-57) | < 0.0001 |
| Japan | 18 | 49 (31-72) | 0.0001 |
| South Korea | 12 | 35 (21-53) | 0.0002 |
| United States | 9 | 29 (17-44) | 0.0003 |
| Cambodia | 1 | 9 (3-16) | 0.0027 |
| Macao, China | 6 | 18 (9-29) | 0.0035 |
| Vietnam | 7 | 17 (9-28) | 0.0110 |
| United Kingdom | 2 | 7 (2-14) | 0.0284 |
| Canada | 2 | 6 (2-13) | 0.0468 |

**Figure 1:**
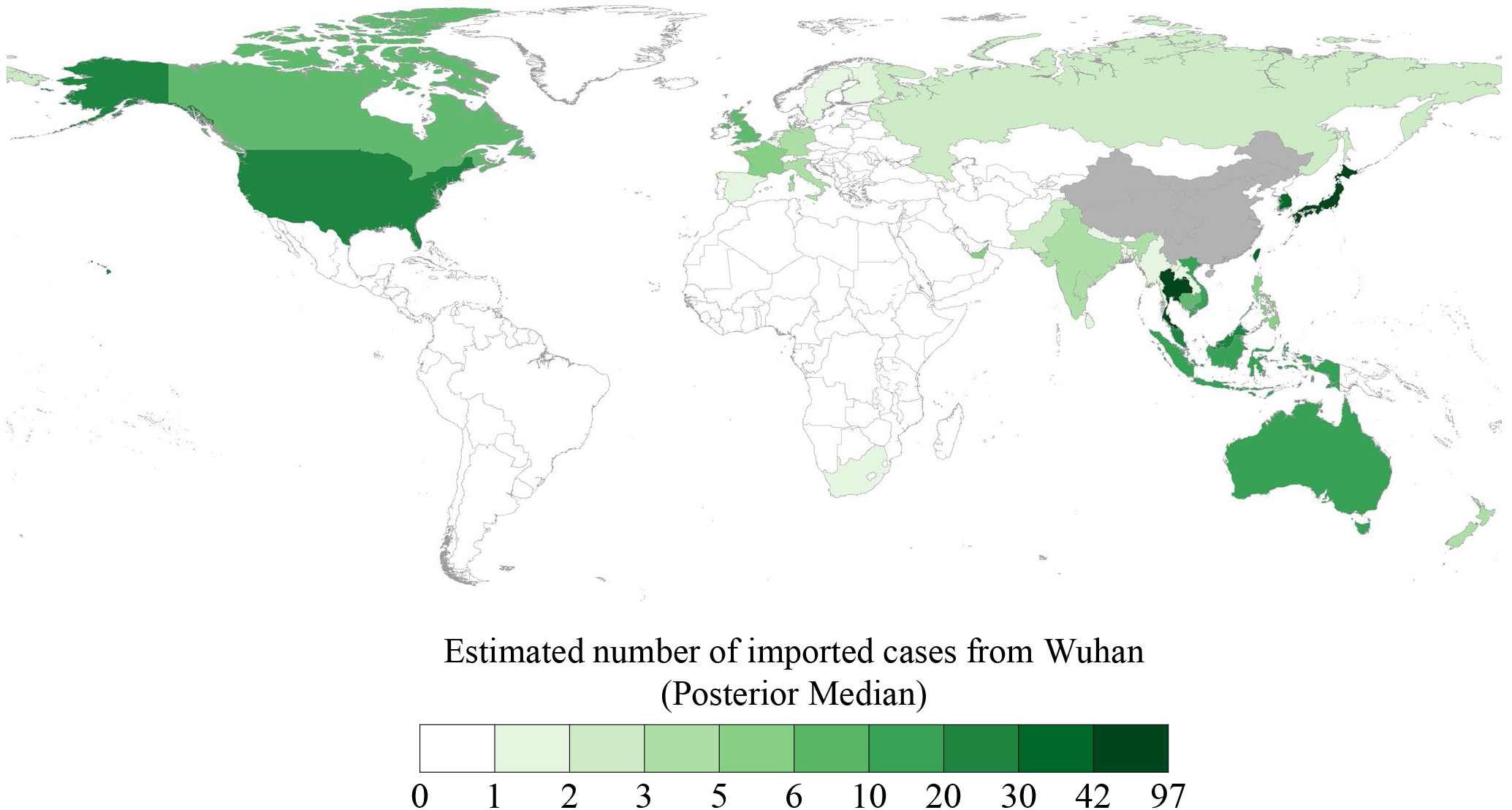
Posterior median estimate of the number of COVID-19 cases imported from Wuhan, for each country or territory outside mainland China. For the countries having the top 10 highest ratios between the reported number of imported cases and the volume of passengers from Wuhan (used as the top reporting rate in our analysis), the reported case counts were shown instead.

If we assume each country has immediately isolated all the *reported* cases imported from Wuhan (and therefore truncated transmission), but not isolated the extra cases we estimated and that no extra control measures are put in place, we estimate that the chance that local transmission would occur and sustain for at least three generations exceeds 50% for a total of 16 countries or territories, including four outside Asia: Australia, Canada, United Kingdom, and United States (Table 2). In a second scenario, where we assume that 95% of all the imported cases from Wuhan were immediately isolated, the estimated local outbreak risk reduces substantially, with only Thailand having a local outbreak risk estimate greater than 50%. Still, the estimated risk of local transmission sustaining for at least three generations is not negligible (> 20%) in many other countries or territories, including Japan, Taiwan, Hong Kong, South Korea, United States, Malaysia, and Singapore (Table 2 for the countries or territories having a local outbreak risk estimate greater than 50% in the main analysis, and Figure 2 & Table S2 for results obtained for all countries or territories outside of mainland China). For countries where we estimate a large number of unreported cases, the local outbreak risk is ranked higher in the first scenario than the second scenario, as the extra estimated cases not being detected means a higher risk of onward transmission compared to countries where we estimate higher detection (Table 2).

**Table 2:**
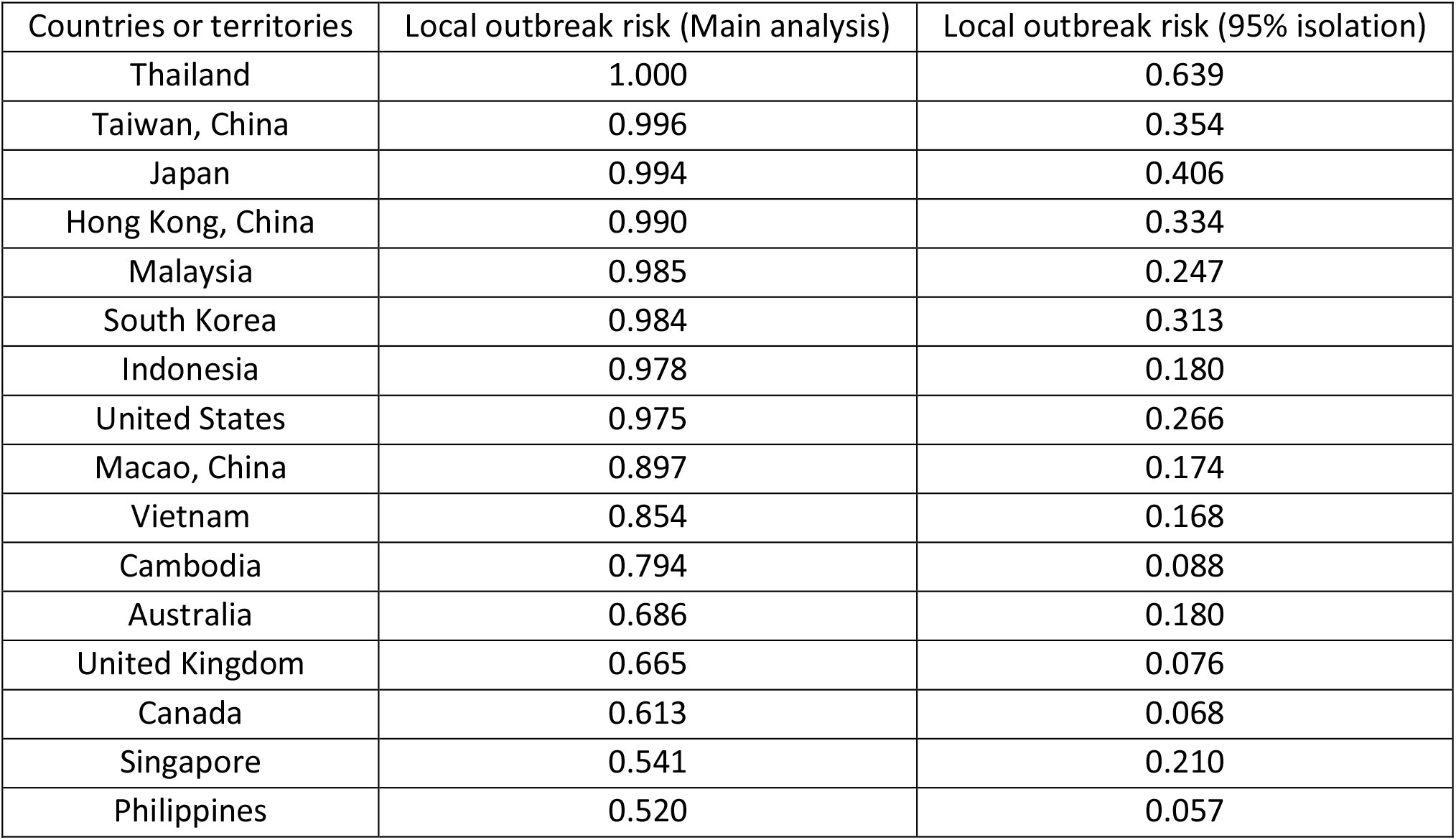
Countries or territories having a local outbreak risk greater than 50% in the main analysis (i.e. assuming immediate isolation of all the *reported* cases imported from Wuhan). In an alternative scenario, we assumed that each country or territory was able to isolate 95% of all the cases imported from Wuhan, and the risk of local outbreak was re-computed (shown in the last column).

**Figure 2:**
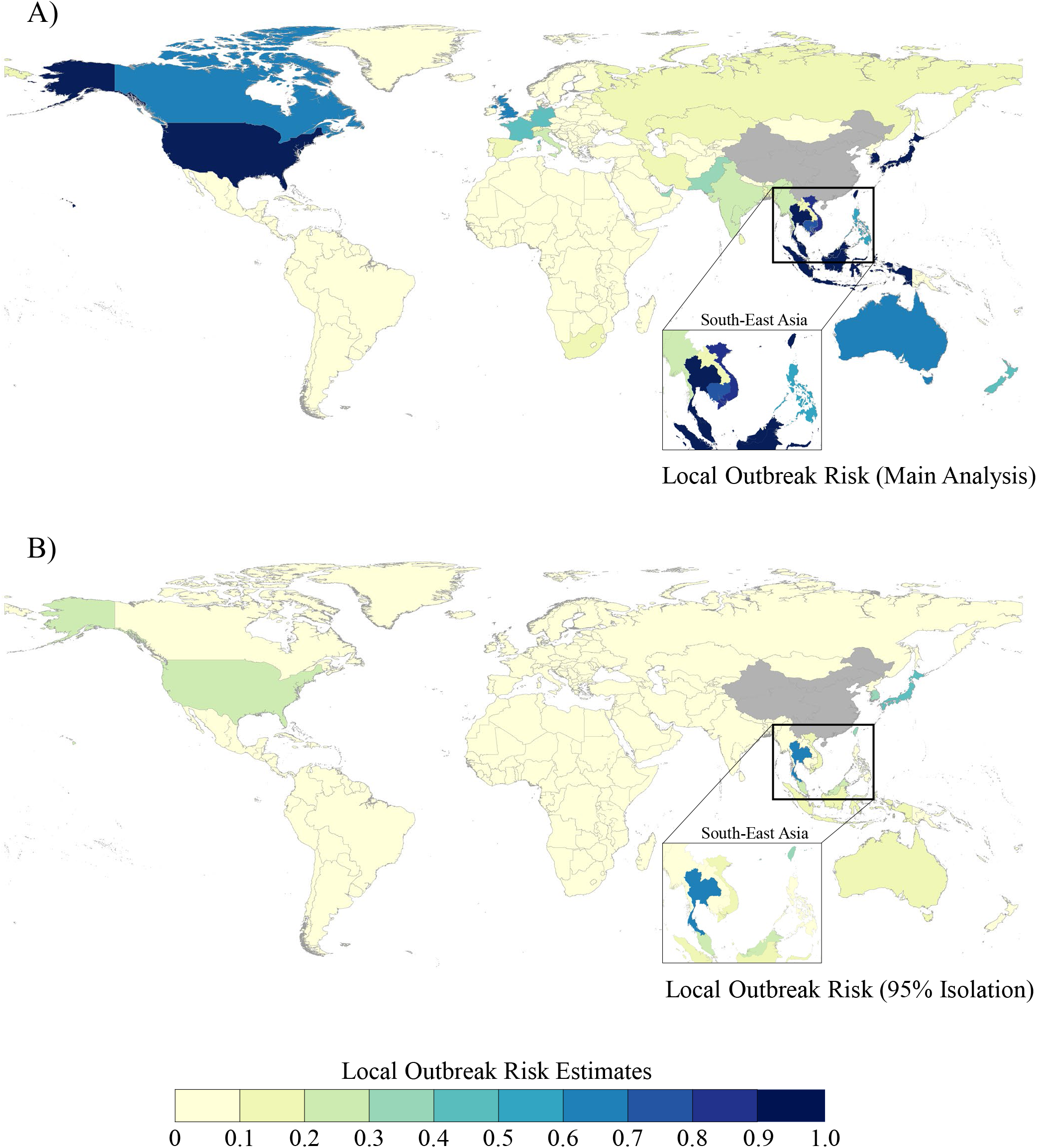
Estimated probability that a local outbreak will occur and sustain for at least 3 generations following the importation from Wuhan (“local outbreak risk”) for each country or territory outside mainland China: (A) Assuming immediate isolation of all the reported cases imported from Wuhan; (B) Assuming immediate isolation of 95% of all the imported cases from Wuhan occurs.

For each recipient country or territory outside mainland China, we assess the relative potential of COVID-19 introduction from each donor country that has a local outbreak risk estimate exceeding 50%. For example, Hong Kong, Singapore, and Australia are found to have the highest COVID-19 introduction index estimates when we consider the United Kingdom as the recipient country. Using the United Kingdom, United States, South Korea, and South Africa as recipient country examples, we highlight the top donor countries or territories based on the COVID-19 introduction index estimate for each of these countries (Figure 3). The COVID-19 introduction index estimates for all pairs of donor-recipient countries create a network of ranked possible importation links between countries outside mainland China (Table S3A). The total COVID-19 introduction index estimate from all the potential donor countries is highest for Taiwan followed by Japan (full list of estimates shown in Table S3B).

**Figure 3:**
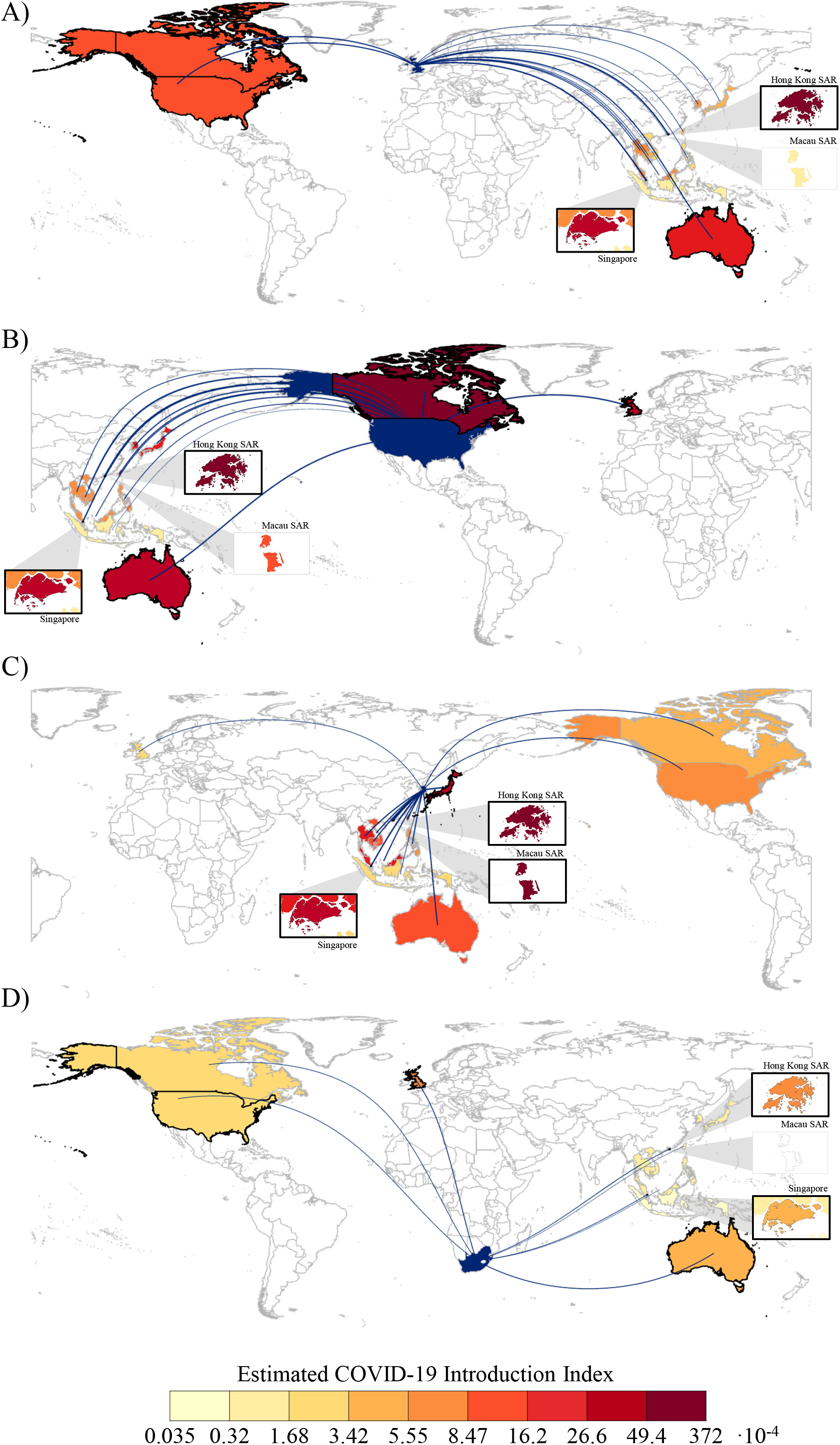
Estimated COVID-19 introduction index between each donor country and (A) United Kingdom, (B) United States, (C) South Korea, and (D) South Africa as the recipient countries. Donor countries refer to countries or territories outside mainland China with a local outbreak risk estimate exceeding 50% in the main analysis. The width of each flight route denotes connectivity between the donor and recipient. Countries with thick black borders or are in a bounding box with a thick black border denote that they are within the top 5 donors in terms of their COVID-19 introduction index estimates for that recipient country.

## Discussion

This study quantifies, with uncertainty, the estimated number of cases imported from Wuhan to countries outside mainland China. As we use information from the places with the highest ratio of the reported case count to the volume of passengers from Wuhan, we are only estimating the number of importations if these places with the highest ratio are capturing all imported cases. Therefore our estimates should be viewed as a lower bound on the number of imported infections, given possible mild and asymptomatic infections. Even given this, we estimate that Indonesia—which has reported 0 cases to date—would be expected to have more cases than this, in line with estimates from Lipsitch et al. ^6^. In addition, in places that have identified some cases—such as Thailand, Japan, South Korea, Taiwan and Hong Kong, the US and Malaysia—we estimate that the imported number of cases from Wuhan is even higher than those reported. In some of these places, testing may be already increasing, but if not, our results would suggest that these places should be targets for increased screening and testing for SARS-CoV-2.

We next probabilistically determine the risk of local transmission within countries outside mainland China. There is of course currently great uncertainty in these estimates, however some of the countries we estimate with the highest probability have indeed reported (limited) local transmission such as Thailand, the US, Singapore, Taiwan, Japan and the South Korea ^18^. This information was not used to generate our estimates, but provides some validation for the model. In countries such as Singapore, intensive contact tracing of cases and testing of all pneumonia cases was implemented^19^, increasing the likelihood of finding community transmission. The impact of control measures after initial detected cases is not included in our model, but may alter the risk of onward transmission and should be considered in further iterations of the model as in other modelling work considering impact interventions under different transmission scenarios.^20,21^ Other countries that we estimate to be at high risk of local transmission, but that have not yet detected transmission ^18^ include Indonesia, Cambodia, Canada and the Philippines. For all these places, our results suggest there could be a consideration of expanding testing of pneumonia or influenza like illness cases beyond those who have currently travelled, to find community transmission as soon as possible after it occurs, as has been ongoing in Singapore^19^ and has been recommended recently in some areas of the US ^22^.

We estimate that in the first wave of inter-country spread not including mainland China the countries at highest risk are still mainly within Asia, the Pacific, North America and Europe. The first importations of cases to and from countries outside mainland China have already occurred, including from Singapore to the UK, ^23^ and Thailand, Japan and Singapore to the South Korea.^12^ Singapore was second highest on our UK donor risk result list after Hong Kong and Japan, Singapore and Thailand were numbers 4, 5 and 6 on the South Korea’s list after Macao, Hong Kong and Taiwan. Given our results and these observations, it would seem prudent for planning to consider the scenario in which the number of countries that may be both recipient and donors of importations will be increasing. Once transmission is confirmed in a possible donor country, our estimates of the risk transmission index are no longer needed, and the number of reported cases and the volume of traffic between countries are what will become important (see Table S3). However it must be considered that transmission may not always be detected quickly in all possible donor countries so estimates of the risk of transmission in donor countries may remain useful for planning. Though many of the countries at risk of second wave importations are similar to those at risk from Wuhan, there are some countries that we did not estimate as having the highest numbers of importations from

Wuhan, but become at increased risk as we consider donor countries outside mainland China. These include India, New Zealand, Spain and Mexico, as they have greater links to countries outside mainland China than to Wuhan, China. With the first imported case in Africa recently reported in Egypt ^24^ (which was estimated to have the highest link with mainland China by Lai et al.^8^,) we must assess where we are in the timeline of the outbreak with respect to importations to Africa, and including the risk of importation from not only mainland China could be important here.

Not included in our analysis currently is the risk of importation from places in China outside Wuhan. There have yet not been enough importations from outside Wuhan to use our current method, and there is currently much uncertainty on the numbers of cases in China to use this data. However as this changes, methods using both types of data could be considered here. The links between other places in China may follow similar patterns to Wuhan and therefore our risk of countries vulnerable for outbreaks and sourcing onward transmission may be similar, but there may be places that are differentially connected to Wuhan and other places in China and so will become at increased risk as importations from China continue. In addition, flights from other places in China to different countries will be truncated at different times for different countries depending on if and when countries halted incoming flights from China. We also currently don’t consider travel across land or sea borders in our analysis.

There are other limitations to our analysis. We assume that R0 in places outside of Wuhan is similar to that estimated from early transmission in Wuhan, however we do not know how variation in climate, population structure, contact patterns, control measures such as contact tracing and quarantine, and other factors may impact transmission. This is an important area for future research based on what is observed in different places and the extension of the analysis to explicitly model transmission in each recipient country can be undertaken as more becomes known.

In summary, we estimate a number of imported cases from Wuhan that were undetected. Given these importations, we estimate a high risk of onward transmission within a number of countries outside mainland China, particularly in those places where cases were not detected, as these undetected cases could not be isolated, and transmission truncated. Given our results of high risk of onward transmission we highlight the importance of wider testing to pick up community transmission as soon as possible after transmission occurs. We also highlight countries that become at increased risk of importation as transmission occurs outside China, and provide results for each country to assess the countries that pose their highest risk of importation.

## Data Availability

Information on the date, location, and travel history of each reported COVID-19 case was synthesized and made publicly available by the nCoV-2019 Data Working Group.

## Notes

### Competing Interest Statement

The authors have declared no competing interest.

### Funding Statement

ARC and BLD were supported by funding from the National Medical Research Councils Centre Grant Program which funds the Singapore Population Health Improvement Centre. The funding source had no role in the study design, data collection, statistical analysis, results interpretation or manuscript writing.

